# More than words: Extra-Sylvian neuroanatomic networks support indirect speech act discourse comprehension in behavioral variant frontotemporal dementia

**DOI:** 10.1101/2020.09.23.20199851

**Authors:** Meghan Healey, Erica Howard, Molly Ungrady, Christopher Olm, Naomi Nevler, David J. Irwin, Murray Grossman

## Abstract

Indirect speech acts— responding “I forgot to wear my watch today” to someone who asked for the time — are ubiquitous in daily conversation, but are understudied in current neurobiological models of language. To comprehend an indirect speech act like this one, listeners must not only decode the lexical-semantic content of the utterance, but also make a pragmatic, bridging inference. This inference allows listeners to derive the speaker’s true, intended meaning—in the above dialogue, for example, that the speaker cannot provide the time. In the present work, we address this major gap by asking non-aphasic patients with behavioral variant frontotemporal dementia (bvFTD, n=21) and brain-damaged controls with amnestic mild cognitive impairment (MCI, n=17) to judge simple question-answer dialogues of the form: “Do you want some cake for dessert?” “I’m on a very strict diet right now”, and relate the results to structural and diffusion MRI. Accuracy and reaction time results demonstrate that subjects with bvFTD, but not MCI, are selectively impaired in indirect relative to direct speech act comprehension, due in part to their social and executive limitations, and performance is related to caregivers’ judgment of communication efficacy. MRI imaging associates the observed impairment in bvFTD to cortical thinning not only in traditional language-associated regions, but also in fronto-parietal regions implicated in social and executive cerebral networks. Finally, diffusion tensor imaging analyses implicate white matter tracts in both dorsal and ventral projection streams, including superior longitudinal fasciculus, frontal aslant, and uncinate fasciculus. These results have strong implications for updated neurobiological models of language, and emphasize a core, language-mediated social disorder in patients with bvFTD.

## 1. INTRODUCTION

*“The chief end of language in communication is to be understood, and words don’t serve well for that end—whether in everyday or in philosophical discourse—when some word fails to arouse in the hearer the idea it stands for in the mind of the speaker*.*”*

--John Locke (1689), “An Essay Concerning Human Understanding”

To paraphrase the famed English philosopher John Locke, human communication does not depend only on decoding the individual meanings of words per se, but rather decoding the speaker’s *idea* represented by those words. Indeed, we do not communicate by volleying single words back and forth in isolation: we communicate through stories, narratives, and conversations (Bell, 2002; Kellas, 2005). This is a critical point that bears significant implications for the experimental methodology adopted by neuroscientists and the theoretical frameworks they endorse in studying language. From this perspective, the methodology we have used to date—studying the neural basis of phonology, morphology, syntax, and semantics—may be too narrowly focused, as these elements alone are often insufficient for comprehension. Instead, when we consider language in an interactive real-world context—as language for *communication*—we recognize that language is polysemous and consequently, listeners must make pragmatic, bridging inferences in order to derive a speaker’s true meaning. In the present study, we address this major gap in traditional neurobiological models of language by focusing on the highly common but often overlooked inferential component of conversational speech.

Indirect speech acts, which are ubiquitous in daily communication, are a canonical example of natural, inferential language. Consider, for instance, if Sally asks Betty, “Do you want some cake for dessert?” and Betty sadly replies, “I’m on a very strict diet right now.” In the given exchange, Sally can easily infer that Betty is declining the cake, even though it is not explicitly stated in her reply. Although indirect speech epitomizes the resource-demanding, socially-constrained nature of language, its processing appears to be both quick and effortless (Clark, 1979). Still unknown, however, is how the brain accomplishes this remarkable feat: what are the cognitive and neural substrates of indirect speech act comprehension?

Historical investigations into the neurobiology of language have typically been limited to studies of speech sounds, words, and sentences. Pioneered by the physicians Paul Broca and Carl Wernicke, the resulting “Wernicke-Lichtheim-Geschwind” (WLG) model emphasizes two primary hubs in left hemisphere peri-Sylvian cortex: the inferior frontal gyrus, specific for language production, and the posterior superior temporal gyrus, specific for language comprehension. While we have now developed a more nuanced understanding of the contributions of these brain regions in supporting language, the WLG model cannot fully account for the complexities of real-world language and communication—how we integrate utterances with prior context so effortlessly, make inferences about speaker meaning, and engage in the rapid back and forth of conversation (Tremblay and Dick, 2016; Hasson et al., 2018).

More recently, we have begun to study natural language discourse—that is, the social use of language, or language for communication. Discourse typically has a supra-sentential structure, and consequently, may require additional neurocognitive resources to disambiguate meaning. Despite its ubiquity in daily language, however, scant attention has been paid to indirect speech acts like the one above—communicative exchanges in which the intended speaker meaning is not directly coded in the lexical-semantic content of the utterance itself (Grice, 1975; Searle, 1975). To address this major gap in natural language use, we study indirect replies, a subtype of indirect speech that boasts several theoretical advantages over previous language domains used to study discourse: 1) they are relatively short and can be tightly controlled, unlike lengthy narratives; 2) their meaning does not become “frozen” due to repeated usage, as with metaphors, idioms, or proverbs; 3) they do not have an affective component, which typically characterizes irony and sarcasm; and 4) they involve an interactive exchange between speakers, which reflects how language is most commonly used. With these factors in mind, we developed a novel, question-answer paradigm manipulating inferential demand—whether a reply is conveyed directly or indirectly.

Unlike previous fMRI studies in healthy adults (Shibata et al., 2011; Basnáková et al., 2013; Jang et al., 2013; Feng et al., 2017), which may be limited due to their correlative nature, we use a patient lesion-model to examine the neurobiological basis of indirect speech. Here, we study patients with behavioral variant frontotemporal dementia (bvFTD), who constitute an ideal cohort to study deficits in “real world” communication (Grossman, 2018). A young-onset neurodegenerative disease, bvFTD is characterized by changes in social comportment, personality, and executive function due to disease in frontal and temporal cortices. Importantly, while patients are grossly non-aphasic, they may show deficits at the discourse level of language. Previous research thus has demonstrated that bvFTD speech is marked by poor narrative organization and limited appreciation of global meaning, abnormal prosody, simplified grammatical structures, and a reliance on concrete concepts and literal meaning (Ash et al., 2006; Farag et al., 2010; Charles et al., 2014; Cousins et al., 2017; Nevler et al., 2017).

Based on previous work from our laboratory and others, we predict that non-aphasic bvFTD patients will show deficits in indirect speech related in part to disease in brain regions associated with an “extended language network” encompassing social, executive, and language regions (Ferstl et al., 2008). We hypothesize further that critical white matter tracts linking these linguistic and extra-linguistic regions may also be disrupted in bvFTD. While the initial WLG model posited only a single white matter tract for language—the arcuate fasciculus, connecting Broca’s and Wernicke’s areas—more recent work has begun to implicate multiple tracts, including the superior longitudinal fasciculus, inferior longitudinal fasciculus, and uncinate fasciculus (Saur et al., 2008; Friederici, 2015; Vassal et al., 2016). It is these tracts that would permit the traditional language network to interact with extra-Sylvian regions-- namely, the executive control and social brain networks that are hypothesized to play a role in discourse processing. Accordingly, and given that bvFTD is known to show significant WM disease (Agosta et al., 2012), we adopt a multimodal approach and use a combination of high-resolution structural magnetic resonance imaging (sMRI) and diffusion tensor imaging (DTI) to expand our understanding of the neural correlates of real-world communication.

## 2. MATERIALS AND METHODS

### 2.1 Participants

Participants included 21 patients with bvFTD, 17 age and education-matched healthy controls, and 17 brain-damaged controls with amnestic mild cognitive impairment (MCI). See Table 1 for a summary of demographic and clinical characteristics. All patients (bvFTD, MCI) were diagnosed by board-certified neurologists (M.G. and D.J.I.) using published criteria and a consensus procedure (Albert et al., 2011; Rascovsky et al., 2011). As some bvFTD patients may develop language deficits associated with semantic variant primary progressive aphasia (svPPA), any patients with symptomatic evidence of svPPA or a score greater or equal to 1 on the Language Supplement of the Clinical Dementia Rating Scale (CDR) (Knopman et al., 2011) were excluded from the sample population. We note here that we chose MCI as our brain-damaged control group rather than svPPA since we wanted all patients to be non-aphasic and capable of performing the discourse task at a reasonable level of proficiency and without obvious language-related deficits. Alternative causes of cognitive difficulty (e.g. vascular dementia, hydrocephalus, stroke, head trauma, primary psychiatric disorders) were excluded by clinical exam, neuroimaging, CSF, and blood tests. As summarized in Table 1, severity of overall cognitive impairment was assessed in patients using the Mini-Mental State Examination (MMSE). On average, patient scores fell in the mild range. Healthy control subjects were verified through negative self-report of a neurological and psychiatric history and a score of greater than or equal to 28 on MMSE. All subjects were recruited from the Penn Frontotemporal Degeneration Center and gave informed consent according to a protocol approved by the Institutional Review Board at the University of Pennsylvania.

**Table 1.**
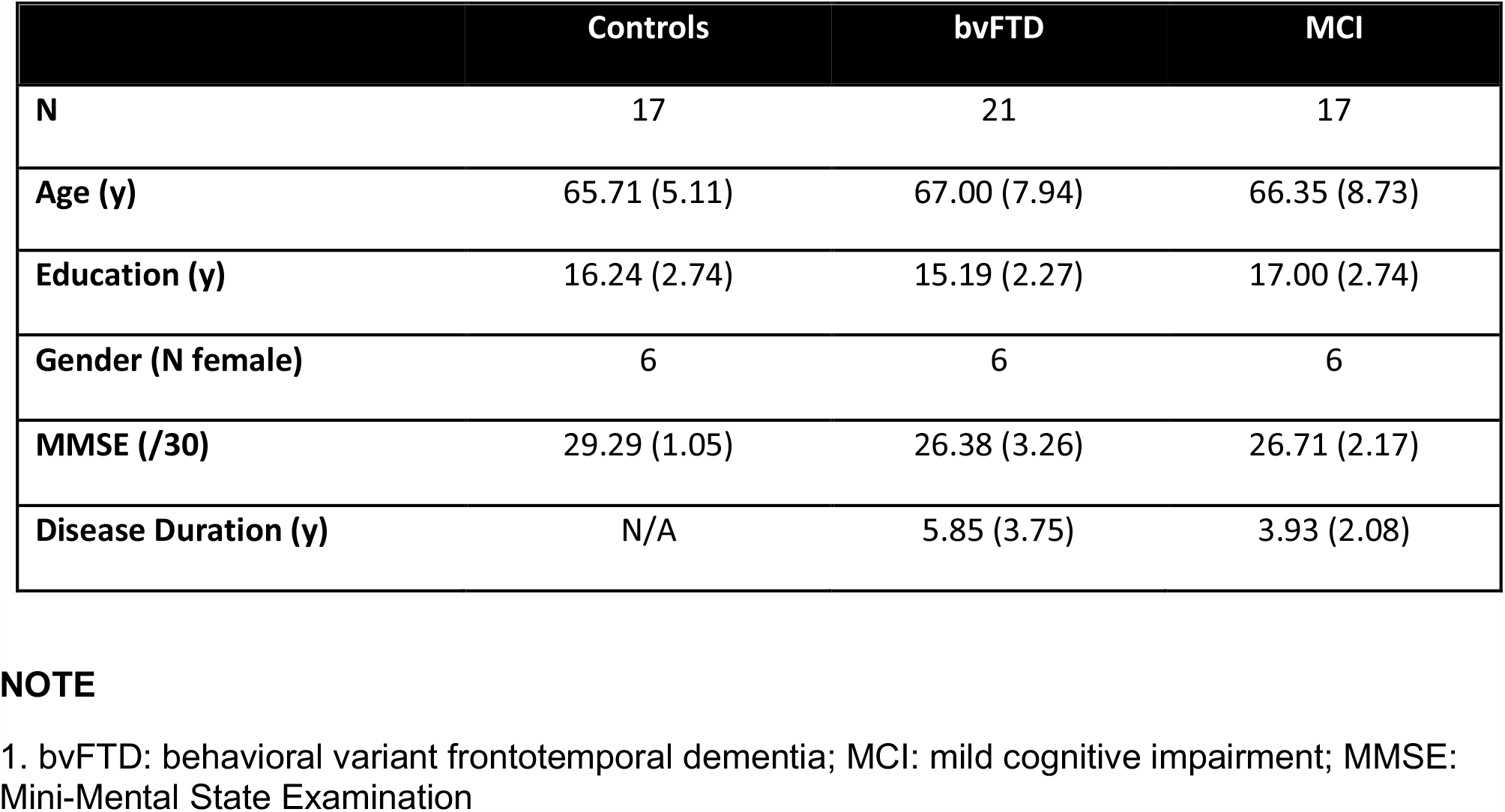
Mean (±SD) of group demographic characteristics^**1**^.

#### 2.2 Experimental Design and Statistical Analyses

The stimulus materials consisted of 120 question-answer dialogues (60 experimental items and 60 filler items of similar structure), summarized in Table 2. They were of the form: “Do you want some cake for dessert?” “I’m on a very strict diet right now”, All questions were polar, such that the expected answer was either “yes” or “no.” Stimuli were presented as printed text in order to avoid any confounds introduced by prosodic cutes inherent in the speech stream or limited working memory.

**Table 2.**
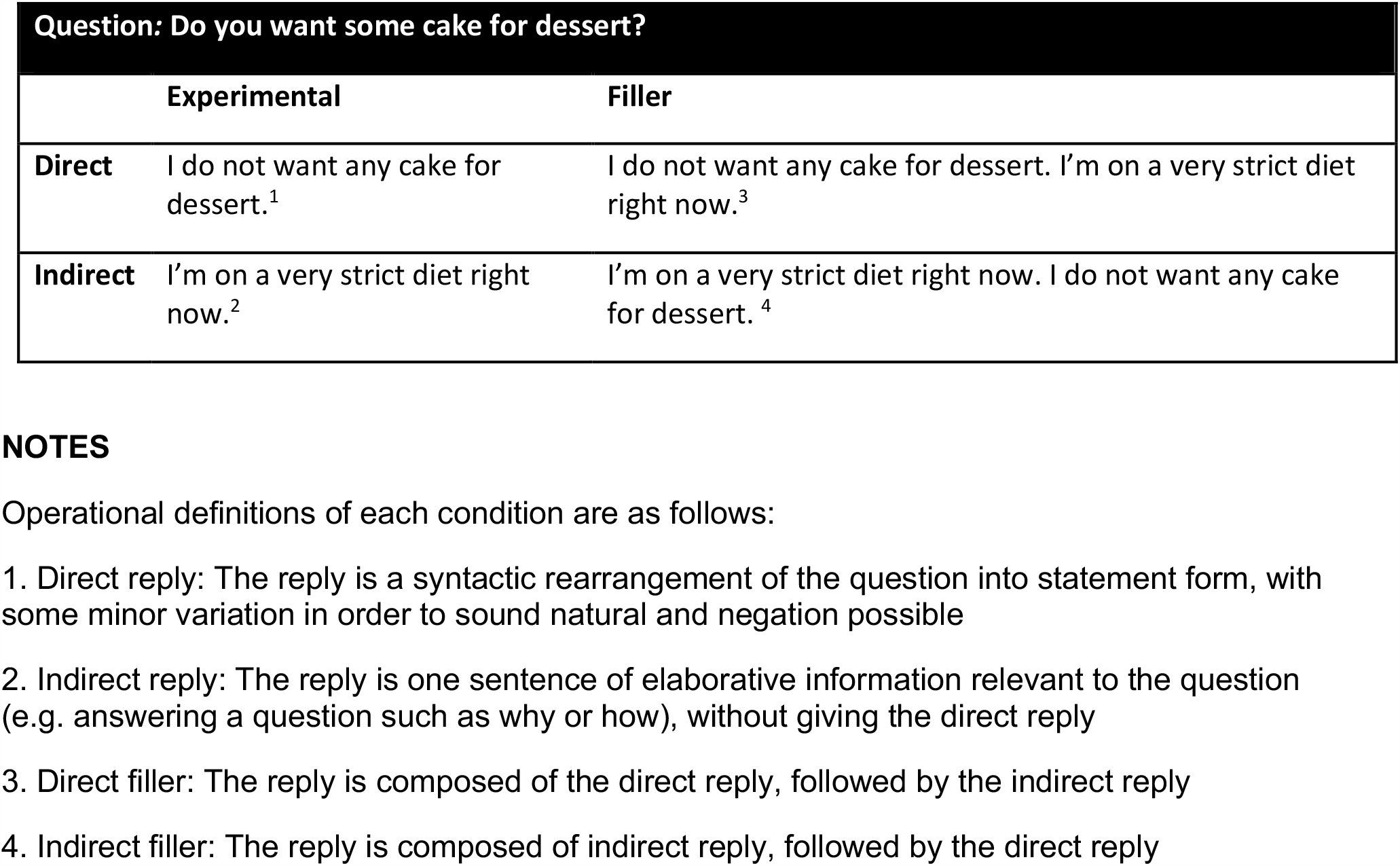
Sample Stimulus Materials.

Each question (n=30) was associated with two different replies, which systematically varied according to inferential demand (direct, indirect). The 60 filler items used the same questions, but presented both the indirect and indirect replies in succession (30 provided the direct reply first, and 30 provided indirect reply first). The filler items will not be discussed further here. Table 2 illustrates each condition and sample stimuli. Note that indirect replies, as operationalized here, are equivalent to Grice’s notion of “conversational implicatures” (Grice, 1975).

Stimuli were carefully constructed to minimize linguistic variation within and across conditions. The direct and indirect items were matched within each item for number of syllables, mean word frequency (Brysbaert and New, 2009), and mean concreteness (Brysbaert et al., 2014). For word frequency and concreteness, a mean score was generated for each sentence by averaging across the individual scores of each content word. This careful matching procedure is meant to ensure that any differences in processing direct and indirect items are due to the manipulation of inferential demand, and not to any differences in other linguistic properties.

Stimulus presentation, timing, and responses were controlled via E-Prime presentation software. On each trial, a fixation cross was presented (3 seconds), followed by the question (3 seconds), and then reply (3 seconds). The question remained on the screen as the reply appeared, in order to reduce any working memory demands. Following each trial, subjects were presented a probe: “Does the reply mean yes or no?” and given 10 seconds to respond via button press. Response accuracy and response time were recorded for each condition. Items were counterbalanced so that half the replies had a positive connotation (i.e. mean “yes”) and half the replies had a negative connotation (i.e. mean “no”). Participants were trained prior to testing and completed 12 practice trials. In total, task administration took approximately one hour.

We assessed performance using two independent metrics: response accuracy and reaction time, as well as two derivative measures: an impairment score and a slowing score. The impairment score, which was meant to quantify a patient’s degree of impairment in indirect speech processing specifically, was calculated by subtracting accuracy in the direct condition from accuracy in the indirect condition within each individual subject (impairment score per subject = indirect accuracy – direct accuracy). The slowing score is an analogous measure for reaction time (slowing score = indirect reaction time -- direct reaction time). All analyses used non-parametric statistics as the data were not normally distributed according to Shapiro-Wilks tests. Between-group comparisons were performed with Mann-Whitney tests, and within-group comparisons with Wilcoxon tests. Correlations were calculated using the Spearman method. All statistical analyses were performed in R (https://cran.r-project.org/).

Prior to data collection, stimulus validity was confirmed via pre-testing. In a norming study, healthy, young adult subjects (n=10) were asked to read each dialogue and respond to a series of question via button press. As in the main experiment, subjects were first asked to indicate if the reply meant “yes” or “no”. Next, subjects were asked to rate how direct the reply sounded and how natural the dialogue sounded, both on a scale of 1 to 5 (where 1= very direct/natural and 5 = very indirect/unnatural). Overall, subjects performed at ceiling, with a mean (+S.D.) accuracy of 97.87% (+0.05) across all items. Furthermore, there was no significant difference for accuracy [direct = 96.88(+0.02), indirect=98.63(+0.01); p=0.07] or naturalness [direct = 1.45(+0.64), indirect = 1.87 (+0.51); p=0.12], in the direct and indirect conditions. Importantly, there was a significant difference between stimuli in terms of the directness rating [direct = 1.19(+0.13), indirect = 3.50 (+0.98); p=0.00003].

#### Neuropsychological Battery

In order to assess the potential contribution of linguistic and non-linguistic cognitive processes to speech act comprehension, both bvFTD and MCI patient groups were also administered a comprehensive neuropsychological battery. Language was assessed with 3 measures, each representing a different level of language processing. Phonological awareness was assessed with the Repetition score from the Philadelphia Brief Assessment of Cognition (PBAC) (Libon et al., 2011), and semantic knowledge with the Multi-Lingual Naming Test (MINT) (Gollan et al., 2012). Finally, grammatical comprehension was assessed using a two-alternative forced-choice sentence-picture matching task, which yields a ratio score comparing comprehension of object-relative sentences to subject-relative sentences (Charles et al., 2014).

Next, executive function was assessed with backward digit span (BDS) (Wechsler, 1997), a test of working memory which requires subjects to repeat an orally presented sequence of numbers in reverse order, and Trailmaking Test B (TMT) (Reitan, 1958), a test of mental flexibility in which subjects must connect a series of dots in ascending order, alternating between letters (A-K) and numbers (1-12). The time to complete Trailmaking Test B (in seconds) was normalized to each subjects time to complete Trailmaking Test A (in which only numbers are presented and there is no switching involved), in order to control for any potential motor differences across subjects.

Social cognition was assessed with the Social Norms Questionnaire (SNQ), a 22-item questionnaire probing social knowledge and an individual’s ability to use context to decide when a behavior is or is not socially appropriate (Panchal et al., 2015). A higher score on the SNQ indicates greater knowledge of social norms. Scoring of the SNQ also yields two subscores: an “Overadhere” score, which refers to the endorsement of socially appropriate behavior as inappropriate (e.g. wearing the same shirt twice in 2 weeks), and a “Break” score, which refers to endorsement of a socially inappropriate behavior as appropriate (e.g. hugging a stranger without asking first). A caregiver informant also completed the Perception of Conversation Index (PCI). Section 1 of the questionnaire assesses caregiver perception of conversational difficulties in patients and includes questions such as “Has difficulty with telephone conversations,” and “Mixes-up the details while telling a story” (Orange et al., 2009; Savundranayagam and Orange, 2011).

We also collected measures in two unrelated cognitive domains to serve as negative controls: visuospatial functioning and episodic memory. Both of these abilities are typically relatively relatively preserved in bvFTD. To assess visuospatial functioning, we used the “copy” measure of Rey-Osterreith Complex Figure Test (Libon et al., 2011), in which a subject must copy a complicated geometric line drawing freehand, and Judgment of Line Orientation (JOLO), in which subjects to match an angled line to one of 11 lines that are arranged in a semicircle (Benton et al., 1983). Finally, to assess episodic memory, we used the “recall” measure of the Rey-Osterreith Complex Figure Test (Libon et al., 2011), where a subject must draw the same complicated line drawing from memory, after a delay. We also assessed episodic memory with Philadelphia Verbal Learning Test (Libon et al., 1996), which is a 9-item list-learning task modeled after the California Verbal Learning Test. The number of correct items recalled on Trial 7 was used as the dependent variable here.

### 2.3 Structural Imaging: Methods and Analysis

#### 2.3.1 Image Acquisition

High-resolution volumetric T1-weighted structural magnetic resonance imaging (MRI) was collected for 19 bvFTD patients and an independent cohort of 25 healthy age- and education-matched controls from the surrounding community (mean age = 67.23 (±7.46), p = 0.37; mean education = 15.88 (±2.19) p=0.22). These controls were used to define an average template brain of comparable age that can be used to identify regions of significant gray matter disease in patients, on a voxel by voxel basis. A T1 image was not available for two patients with bvFTD due to contraindications and safety concerns, including claustrophobia and metal in the body (i.e pacemaker). MRI volumes were acquired using a magnetization prepared rapid acquisition with gradient echo (MPRAGE) sequence from a SIEMENS 3.0T Tim Trio scanner using an axially acquired protocol with the following acquisition parameters: repetition time (TR)=1620 ms; echo time (TE)=3.87 ms; slice thickness=1.0 mm; flip angle=15°; matrix=192×256, 160 slices, and in-plane resolution= 0.9766×0.9766 mm^2^. Whole-brain MRI volumes were preprocessed using Advanced Normalization Tools (https://github.com/ANTsX/ANTs) using the state-of-the-art antsCorticalThickness pipeline described previously (Avants et al., 2008; Klein et al., 2010; Tustison et al., 2014)(Avants et al., 2008; Klein et al., 2010; Tustison et al., 2014). Briefly, processing begins by deforming each individual dataset into a standard local template space that uses a canonical stereotactic coordinate system, generated using a subset of images from the open access series of imaging studies dataset (OASIS) (Marcus et al., 2010). ANTs then applies a highly accurate registration algorithm using symmetric and topology-preserving diffeomorphic deformations, which minimize bias to the reference space while still capturing the deformation necessary to aggregate images in common space. The ANTs Atropos tool uses template-based priors to segment images into six tissue classes (cortex, white matter, CSF, subcortical gray structures, brainstem, and cerebellum) and generate corresponding probability maps. Voxelwise cortical thickness is finally measured in millimeters (mm). Resulting images are warped into Montreal Neurological Institute (MNI) space, smoothed using a 2 sigma smoothing kernel, and downsampled to 2mm isotropic voxels.

#### 2.3.2 Voxel-wise Analyses

To define areas of significant cortical thinning in bvFTD, non-parametric, permutation-based imaging analyses were performed with threshold-free cluster enhancement (Smith and Nichols, 2009) and the randomise tool in FSL (http://fsl.fmrib.ox.ac.uk/fsl/fslwiki). Briefly, permutation-based t-tests evaluate a true assignment of cortical thickness values across groups (signal) relative to many (e.g. 10,000) random assignments (noise). Accordingly, permutation-based statistical testing is robust to concerns regarding multiple comparisons and preferred over traditional methods using parametric-based t-tests as permutation testing effectively controls for false positives (Winkler et al., 2014). Cortical thickness was compared in patients relative to the independent cohort of 25 healthy controls described above and restricted to an explicit mask of high probability cortex (>0.4). We report clusters that survived a statistical threshold of p<0.01, correcting for multiple comparisons using the family wise error (FWE) rate relative to 10,000 random permutations. Results were projected onto the Conte69 surface-based atlas using Connectome Workbench (http://www.humanconnectome.org/software/connectome-workbench.html).

To relate behavioral performance to regions of significant cortical thinning, we fit linear regression models with the randomise tool of FSL and the impairment score as a covariate. Permutations were run exhaustively up to a maximum of 10,000 for each analysis. To constrain our interpretation to areas of known disease, we restricted our regression analyses to an explicit mask containing voxels of significant cortical thinning, as defined by the group comparison described above. Results outside these regions of known disease would be difficult to interpret since they could be attributed to a variety of individual differences unrelated to disease per se (e.g. healthy aging, genetic variation, etc.). For the regression analyses, we report clusters with a minimum of 20 adjacent voxels and surviving a height threshold of p<0.005, which is recommended for optimal balance of Type I and Type II error rates (Lieberman and Cunningham, 2009). Results were projected onto slices using MRIcron software (Rorden and Brett, 2000).

#### 2.3.2 ROI Analyses

Next, we conducted a series of whole-brain region-of-interest (ROI) analyses in order to specifically test our hypothesis that indirect reply comprehension involves the interaction of multiple brain networks: the core language network, the theory-of-mind/social network, and the multiple-demand network/executive network. Using publicly available software (https://github.com/ftdc-picsl/QuANTs/tree/master/R), we extracted mean cortical thickness values for each of the 3 networks for each subject. Each network ROI (see below for network ROI definitions) was warped from MNI space to the subject’s native T1 space prior to extracting the estimates of cortical thickness (mm). To demonstrate specificity of our predicted relationship, we similarly extracted cortical thickness estimates from the sensorimotor network to use as a negative control.

The language network ROI was constructed by summing 4 language ROIs identified by (Fedorenko et al., 2010, 2013), who used a language localizer contrasting reading sentences to reading lists of unconnected, but pronounceable words. The final ROIs, which included left IFG, IFG (pars orbitalis), anterior temporal lobe, and posterior temporal lobe, were created from a probabilistic overlap map from 220 healthy participants.

The social network was the sum of 7 ROIs, which were originally constructed by Dufour et al., (2013) and included: dorsomedial prefrontal cortex (dmPFC), middle medial prefrontal cortex (mmPFC), ventromedial prefrontal cortex (vmPFC), precuneus (PC), right superior temporal sulcus (RSTS), right temporoparietal junction (RTPJ), and left temporoparietal junction (LTPJ). The ROIs were developed by contrasting the false belief and false photograph conditions of a standard story-based theory-of-mind task across 462 healthy participants.

The executive ROIs were also adopted from Fedorenko et al. (2013), who contrasted hard and easy versions of a spatial working memory task in 197 healthy participants. For our purposes, we summed only those MDN ROIs overlapping the so-called “fronto-parietal attention network.” The ROIs we selected thus included dorsolateral prefrontal cortex, orbital middle frontal gyrus, bilateral superior parietal lobe, inferior parietal sulcus, and inferior parietal lobule.

Finally, our control sensorimotor network was taken from Shirer et al. (2012), who defined ninety functional ROIs across 14 large-scale resting state brain networks using a classifier with leave-one-out cross-validation. Please see Figure 3 for the anatomic distribution of these brain networks.

### 2.4 Diffusion Tensor Imaging: Procedure and Analysis

White matter tracts play a critical role in network activity by transmitting electrical signals across spatially separate gray matter regions, both within and across hemispheres. Therefore, even when gray matter regions are intact, synchronized network activity can be disrupted if there is damage to the white matter projections connecting gray matter nodes to each other. Because of this possibility, we use diffusion tensor imaging to examine patterns of structural connectivity in bvFTD and build a large-scale, multimodal network underlying speech act comprehension.

Diffusion weighted imaging (DWI) was available for the same 19 bvFTD patients with T1 imaging. A 30-directional DWI sequence was collected using single-shot, spin-echo, diffusion-weighted echo planar imaging (FOV=240 mm; matrix size =128×128; number of slices = 70; voxel size = 1.875×1.875×2.2 mm3, TR = 8100 ms; TE = 83 ms; fat saturation). Thirty volumes with diffusion weight (b=1000 s/mm2) were collected along 30 non-collinear directions, and either one or five volumes without diffusion weight (b=0 s/mm2) were collected per subject. A chi-square test demonstrated that the distribution of subjects with either one or five volumes without diffusion weight did not differ significantly across our groups (χ2=5.4412, p=0.08). We also include a nuisance covariate for sequence in our subsequent analyses.

The diffusion images were processed using ANTs (Tustison et al., 2014) and Camino (Cook et al., 2006). Motion and distortion artifacts were removed using affine co-registration of each diffusion-weighted image to the average of the unweighted (b= 0) images. Diffusion tensors were calculated using a weighted linear least-squares algorithm (Salvador et al., 2005) implemented in Camino. Fractional anisotropy (FA) was computed in each voxel from the DT image, and distortion between the subject’s T1 and DT image was corrected by registering the FA to the T1 image. DTs were then relocated to the local template for statistical analysis by applying the FA-to-T1, T1-to-local template, and local template-to-MNI warps, and tensors were reoriented using the preservation of principal direction algorithm (Alexander et al., 2002). Each participant’s FA image was recomputed from the DT image in MNI152 template space and smoothed using a 2-sigma smoothing kernel.

Like the pipeline for GM analysis, we used the randomise tool in FSL to compare FA in patients relative to the same cohort healthy age-matched controls. The two-sample t-test of patients vs. controls was run with 10,000 permutations and restricted to voxels containing WM based on an explicit mask of high probability WM (minimum FA considered WM = 0.20). We also include a nuisance covariate of no interest for sequence difference (sequences with one versus five volumes without diffusion weight). We report clusters that survived a statistical threshold of p<0.005 and a minimum cluster extent of 200 voxels. Regression analyses then related patient impairment to reduced FA, using a covariate for the indirect impairment score and a nuisance covariate for sequence. These regressions were restricted to the results of the previous analysis—that is, only voxels showing a significant effect of group. As above, we report clusters surviving a height threshold of p<0.005 and a minimum of 20 contiguous voxels.

### 2.5 Data Availability

The data for the study are available from the authors to qualified investigators with appropriate Institutional Review Board approval and Material Transfer Agreement.

## 3 RESULTS

### 3.1 Analysis of Task Performance

Our first objective was to test the hypothesis that inferential demand (i.e. whether a reply was communicated directly versus indirectly) modulates response accuracy in bvFTD. Results are summarized in Figure 1. We find that healthy control subjects performed at ceiling in both direct and indirect conditions, with no significant difference between conditions (W=23.5, p=0.10). Patients with bvFTD, on the other hand, perform significantly worse in the indirect condition than the direct condition (W=132.5, p=0.0008). bvFTD patients were also significantly impaired relative to healthy controls in the indirect condition (W=276.5, p=0.003), but not the direct condition (U=237.5, p=0.07). The null result in the direct condition suggests that segmental language ability, such as the comprehension of single words and sentences, is unlikely to be responsible for the decrement in indirect performance. To confirm the group-level results in bvFTD, we also calculated an “impairment score” by subtracting accuracy in the direct condition from accuracy in the indirect condition within each individual subject (impairment score = indirect accuracy – direct accuracy). Accordingly, more negative scores represent a greater degree of impairment. Results again indicate that bvFTD patients (mean impairment = −0.08 (±0.09)) are significantly more impaired than healthy controls (mean impairment = −0.01 (±0.02), U=273.00, p=0.004). Sixteen of 21 (76.20%) bvFTD patients showed a negative impairment score.

**Figure 1.**
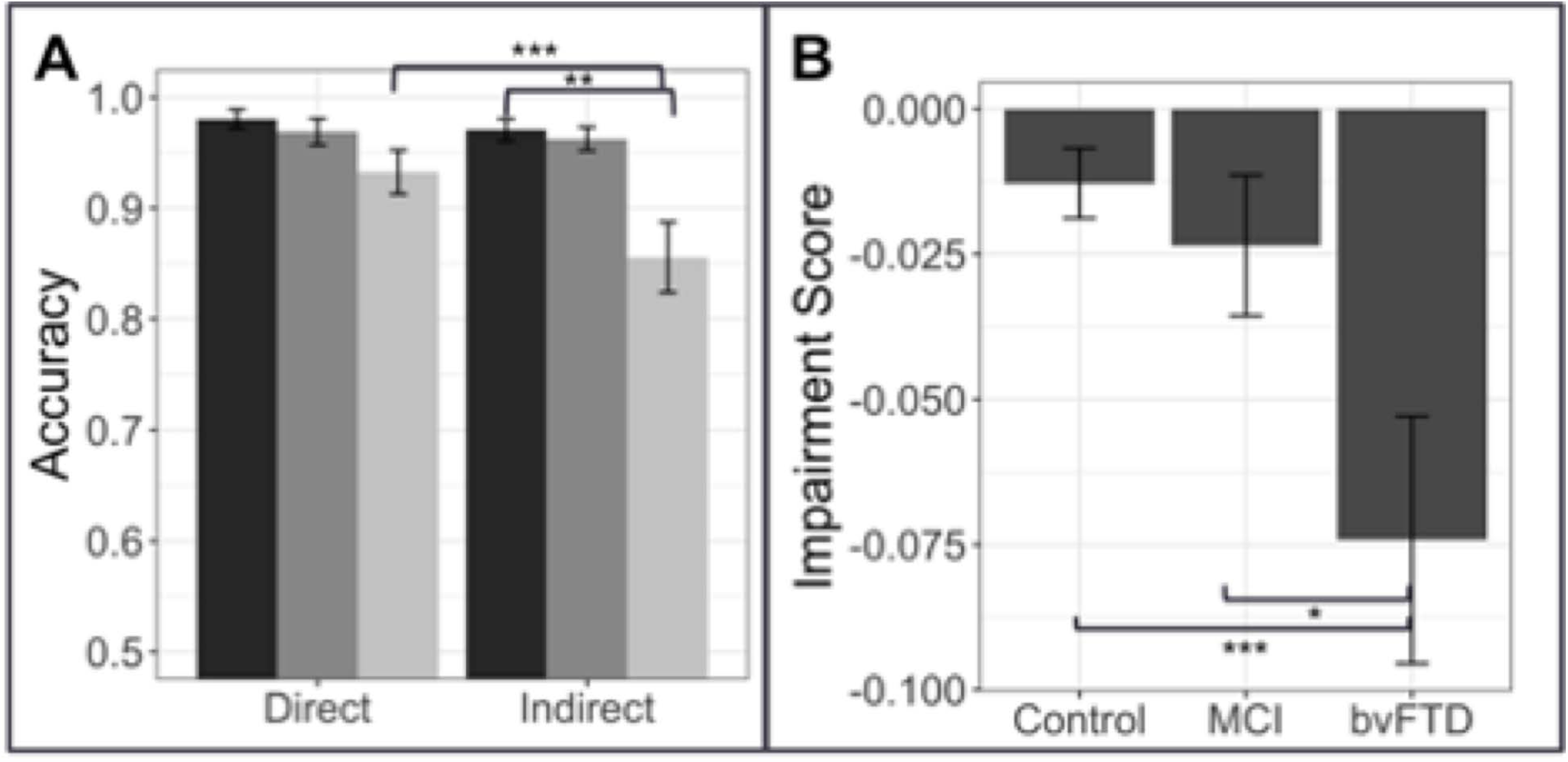
Response Accuracy: Response accuracy in controls, patients with behavioral variant frontotemporal dementia (bvFTD), and patients with amnestic mild cognitive impairment (MCI) in the experimental (short) conditions. A. Mean (±SEM) accuracy in the direct and indirect conditions. Controls are shown in dark gray (left-most bar), bvFTD patients in medium gray (middle bar), and MCI patients in light gray (right bar). B: Mean (±SEM) impairment score (indirect – direct) in the short condition across groups. A more negative impairment score indicates more difficulty with the indirect condition relative to a patient’s individual baseline performance on the direct condition. * indicates significance at p<0.05, ** indicates significance at p<0.01, *** indicates significance at p<0.001.

Patients with MCI show some cognitive decline for their age but remain largely capable of independent day-to-day functioning (Gauthier et al., 2006) and thus represent an appropriate brain-damaged control group to test the specificity of the effect observed in bvFTD. Results in MCI show that, unlike bvFTD patients, MCI patients are not significantly impaired relative to healthy controls in either the direct (U=165.50, p=0.43) or indirect condition (U=170.00, p=0.36). Similarly, their mean impairment score (−0.006 (±0.06)), calculated within each individual, does not differ from that of healthy controls (U=171.00, p=0.35), but does differ from bvFTD (U=110.00, p=0.04) (please see Figure 1). Because bvFTD and MCI patients are matched in terms of global cognition as assessed by the MMSE (U=107.5, p=0.93), it is difficult to attribute the relative impairment observed in bvFTD entirely to an effect of overall cognitive impairment, but is more likely to be associated with the deficits characteristic of bvFTD.

The reaction time data offer converging evidence for our claim that patients with bvFTD are selectively impaired in indirect reply comprehension, relative to both healthy controls and patients with MCI. A Kruskal-Wallis test indicates that there are significant differences across our three groups for reaction time in both the direct condition (χ^2^(2)= 10.97, p=0.001) and the indirect condition (χ^2^(2)=13.75, p=0.001). Upon further analysis, we find that patients with bvFTD are significantly slower to respond to direct replies than healthy controls (U=81.5, p=0.004), but not MCI (U=155.00, p=0.5). More importantly, in the indirect condition, bvFTD are slower to respond than both groups (controls: U=67.5, p=0.001; MCI: U=108.00, p=0.038).These data, however, do not address whether bvFTD patients have slower, non-specific motor reaction times or are more affected by the increased inferential demand characteristic of the indirect condition relative to the two other subject groups. To answer this question, we computed an individualized “slowing score” (slowing score = indirect RT - direct RT), analogous to the impairment score calculated for accuracy. In this case, a positive slowing score means a subject is relatively slower in the indirect condition. We find a significant difference in slowing scores across our three groups (χ^2^ =9.30, p=0.001). Post-hoc testing indicates that patients with bvFTD have significantly larger slowing scores than healthy controls (U=81.5, p=0.005) and MCI (U=99.00, p=0.019) (please see Figure 2). Therefore, the disproportionate slowing for indirect compared to direct stimuli in bvFTD suggests that our observations cannot be easily attributed to simple motor slowing. Moreover, this finding demonstrates that patients with bvFTD do not slow their performance in a strategic effort to improve accuracy. Taken together, our data confirm that patients with bvFTD struggle to process indirect replies during conversation both quickly and accurately.

**Figure 2.**
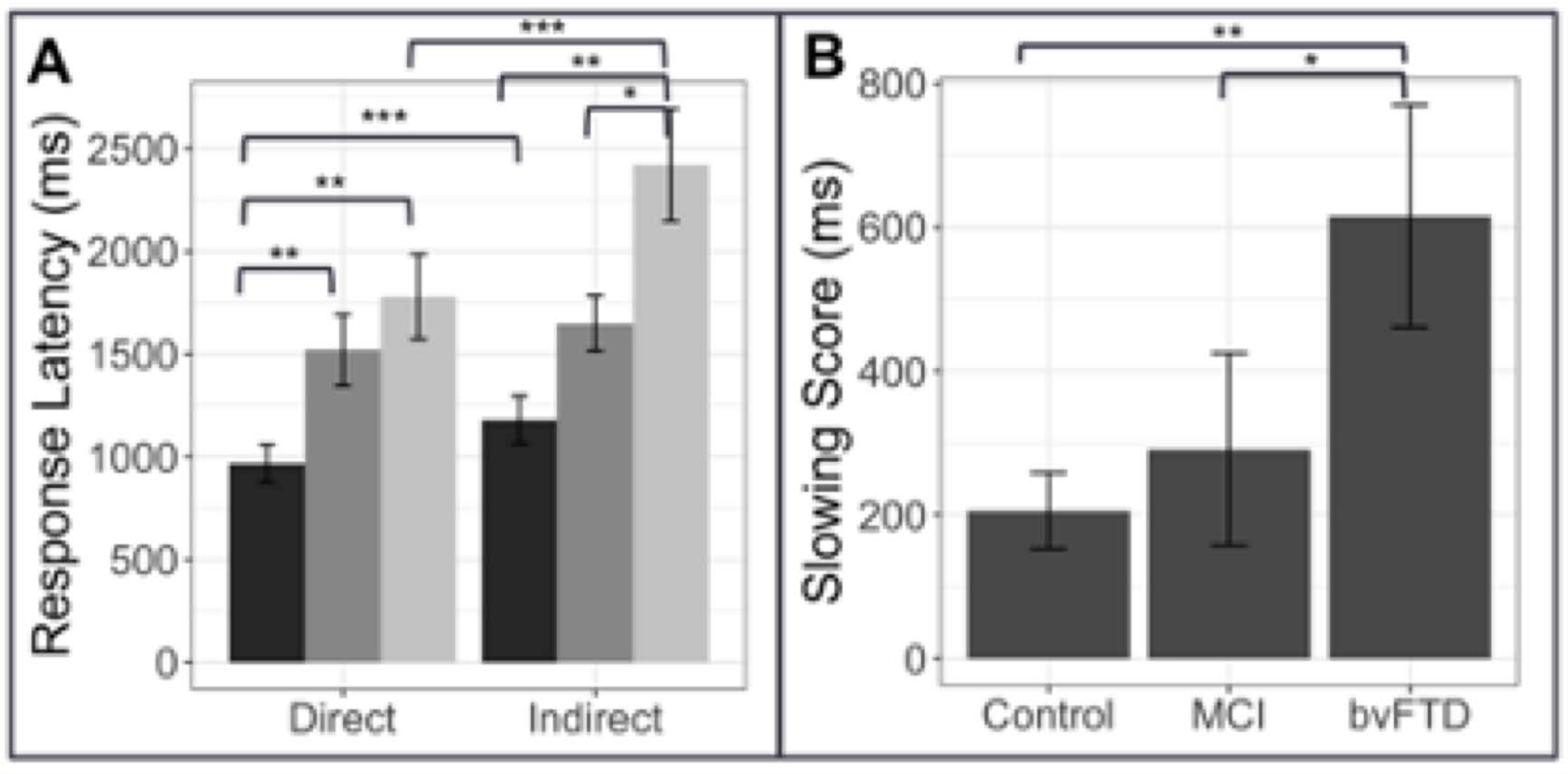
Response latency: Response latency in controls, patients with behavioral variant frontotemporal dementia (bvFTD), and patients with amnestic mild cognitive impairment (MCI). Panel A. Mean (±SE) reaction time in the direct and indirect conditions. Controls are shown in dark gray (leftmost bar), bvFTD patients in medium gray (middle bar), and MCI patients in light gray (right bar). Panel B: Mean (±SE) slowing score in the short condition across groups. A higher slowing score indicates longer reaction times in the indirect condition relative to a patient’s individual baseline performance in the direct condition. * indicates significance at p<0.05, ** indicates significance at p<0.01, *** indicates significance at p<0.001.

**Figure 3.**
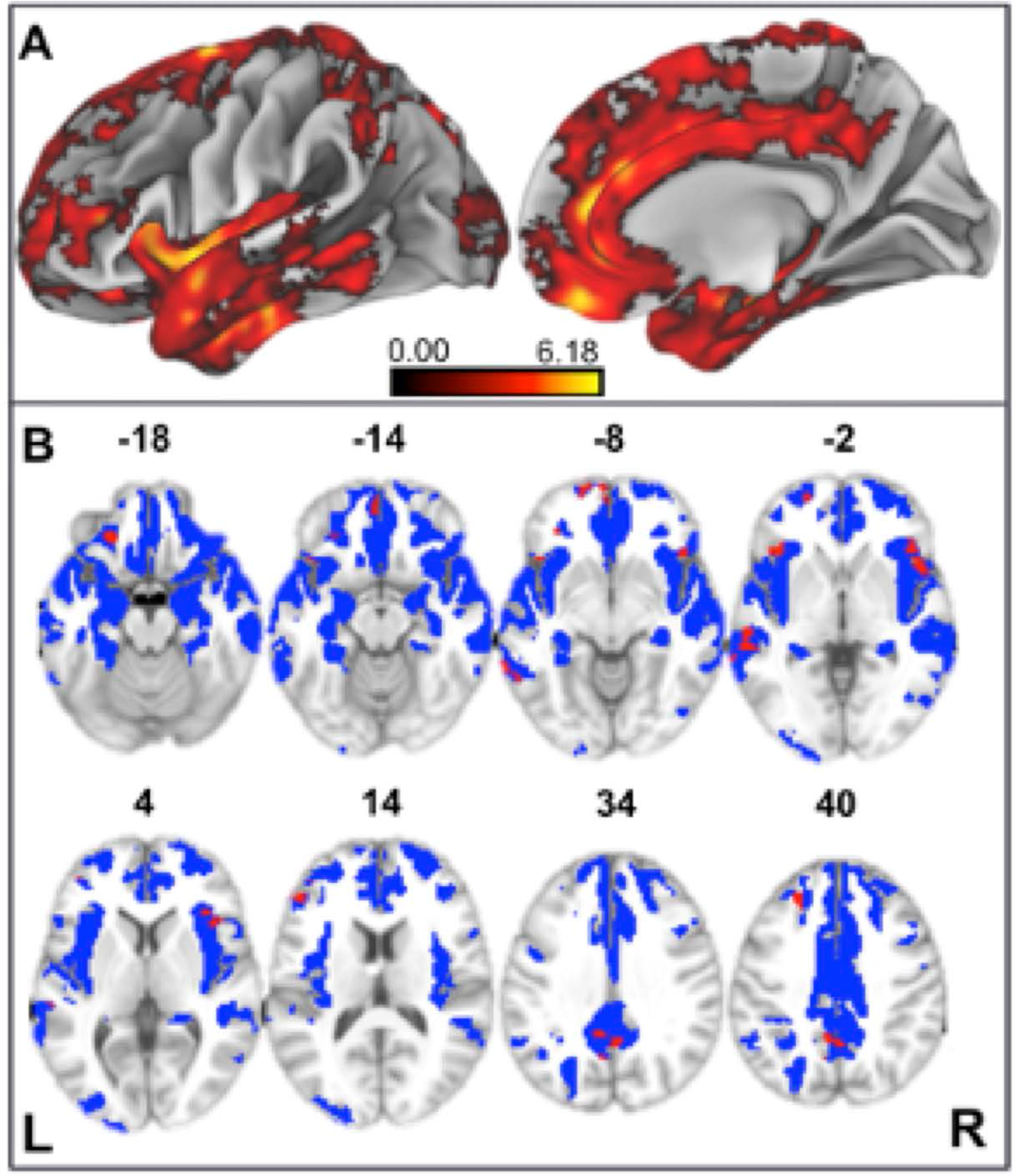
Structural Neuroimaging Results: Panel A: Surface renderings depicting regions of significant cortical thinning in behavioral variant frontotemporal dementia (bvFTD) patients relative to age-matched healthy controls. Heat map intensity refers to t-statistic values. Panel B: Axial slices and z-axis coordinates illustrating regions of significant cortical thinning in bvFTD patients relative to age-matched healthy controls (red and blue regions) and regions of significant cortical thinning associated with indirect comprehension impairment in bvFTD (red areas, only).

### 3.2 Correlational Analyses with Neuropsychological Measures

Next, to examine the cognitive mechanism(s) associated with the observed deficits in bvFTD patients, we administered a broad neuropsychological battery targeting core language skills, executive function, and social cognition that may contribute to inferential comprehension, as well as negative control measures of visuospatial functioning and episodic memory. We used Spearman correlations to relate these independent measures to the indirect-relative-to-direct inferential impairment score.

Our first aim is to demonstrate that indirect speech act comprehension impairment in bvFTD discourse is largely independent of segmental language ability. Consistent with our earlier finding of intact performance in the direct condition, correlation analyses indicate that language ability at both the phonological (i.e. repetition test) and single word levels (i.e. MINT) is not related to inferential impairment (please see Table 3). A measure of grammatical comprehension, however, comparing comprehension of object-relative sentences to subject-relative sentence, is significantly correlated with inferential impairment (rho= 0.52, p=0.04).

**Table 3.**
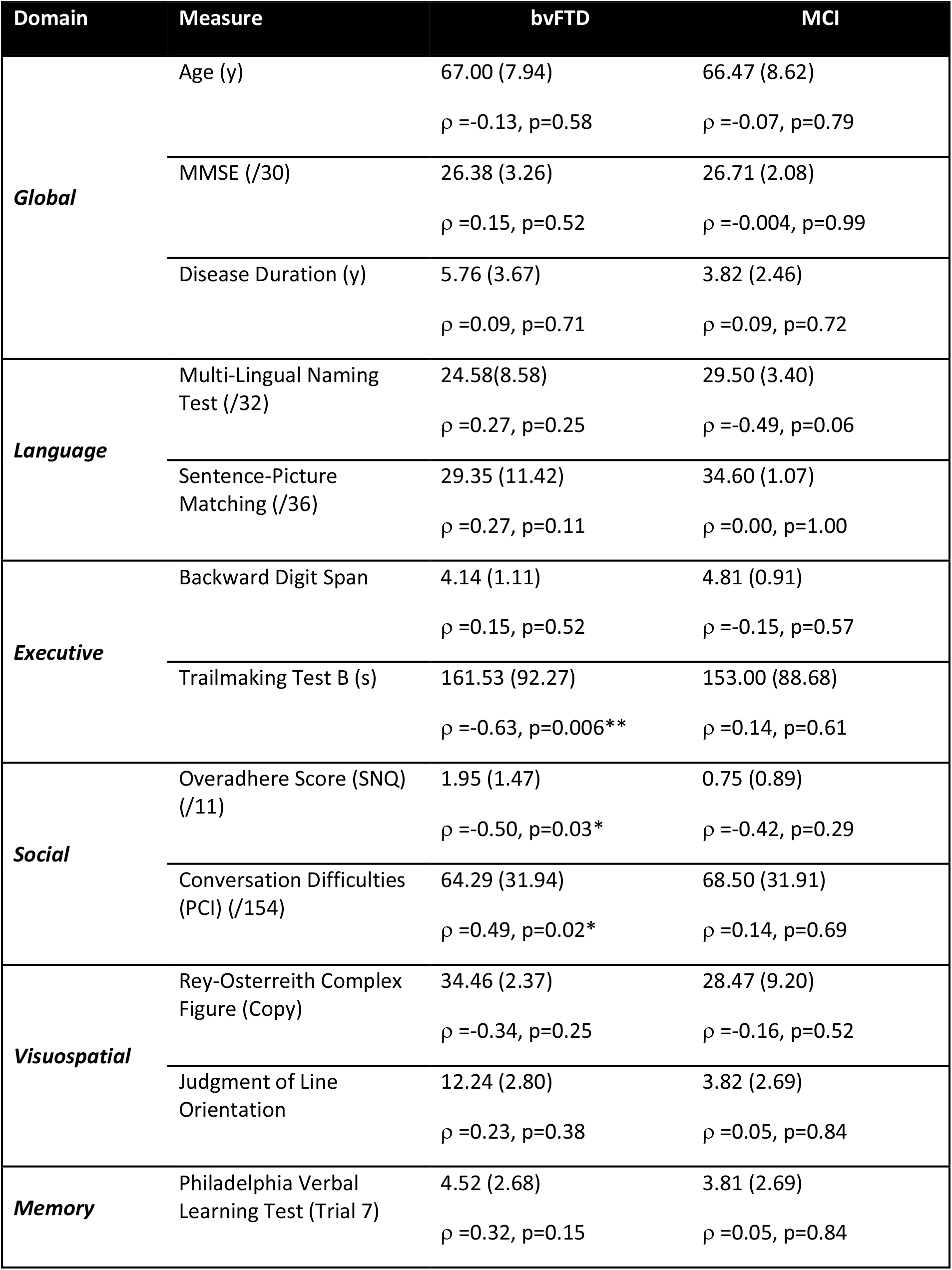
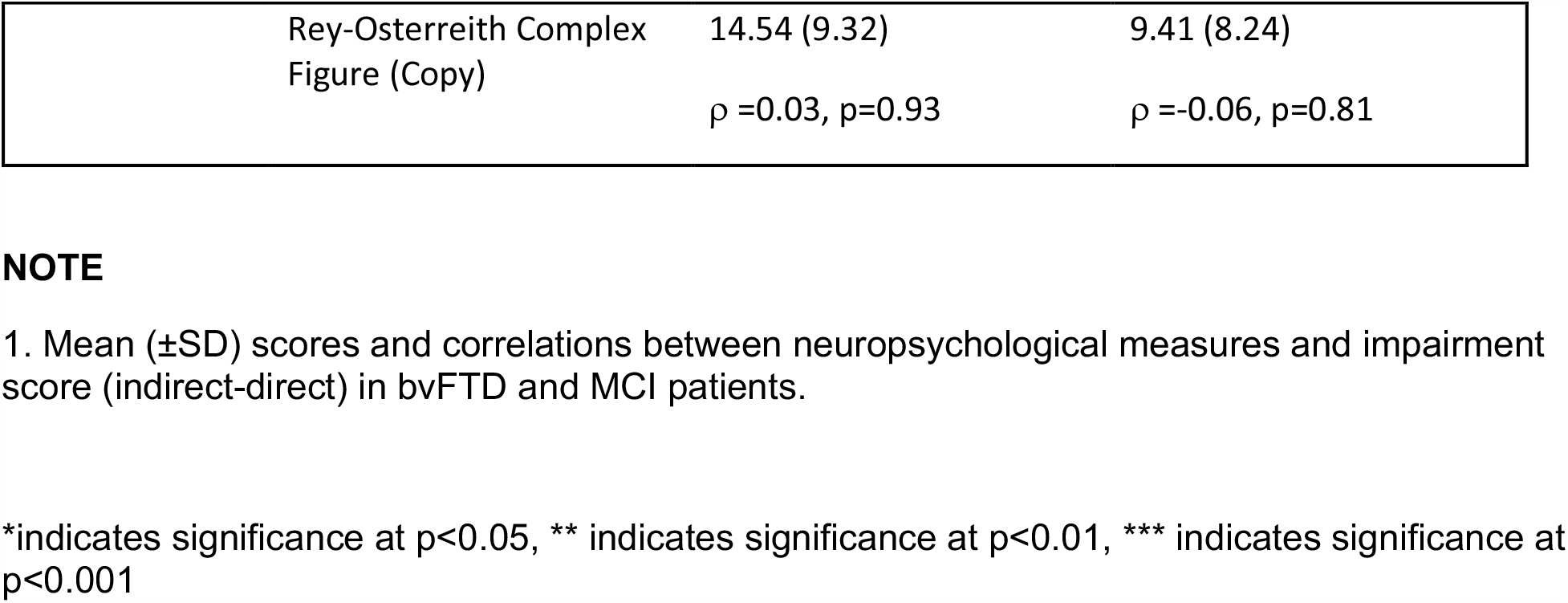
Correlation Results^**1**^.

Next, although patients with bvFTD are known to have deficits in working memory capacity (Kramer et al., 2003; Libon et al., 2007; Baez et al., 2016), we find no relationship between backward digit span and inferential impairment. Other domains of executive function, however, do demonstrate an effect: poor task-switching ability (as indicated by Trailmaking) is correlated with inferential impairment (Trailmaking: rho=-0.63, p=0.006), suggesting a role for mental flexibility in the interpretation of indirect replies.

In the social domain, the impairment score is also positively associated with total score of the SNQ (rho=0.47, p=0.04). Upon further examination, we find that most patients performing worse on the SNQ have a higher Overadhere score than Break score [Overadhere: mean=1.95 (+1.47); Break: mean=1.05 (+1.35)] suggesting that patients who are more rigid in their application of rules to behavior may be similarly rigid in their interpretation of discourse. Finally, we also confirm the construct validity of our indirect speech task by demonstrating that impairment on the task is correlated with real-world conversational difficulties, as assessed by caregivers in the PCI-DAT (rho=0.49, p=0.02).

bvFTD performance in the indirect condition is not related to visuospatial or episodic memory functioning. The same correlation analyses are also performed in patients with MCI in order to test the specificity of the results in bvFTD, and no results in MCI are significant. In sum, we conclude that a relative impairment in understanding indirect speech is specific to bvFTD and related to the social and executive deficits that characterize the disease. More specifically, we implicate the ability to adapt behavior to changing rules and/or contexts in the interpretation of indirect speech.

Based on these initial correlation results, we then used multiple linear regression to predict the impairment score based on three significant and possibly interacting variables, one from each domain: grammatical comprehension, Trailmaking (B-A), and SNQ. A total of 5 different models were tested: all variables as independent (Model 1); all variables interacting (Model 2); and each of the pairwise interactions (the remaining predictor as independent, Models 3-5). Only one model yields a significant regression equation [(Impairment Score ∼ Grammatical Comprehension + Trailmaking * SNQ); F(4,13)=9.346, p=0.006]. The overall model fit is strong, with R^2^=0.84. Both Trailmaking (β=0.009, p=0.004) and SNQ (β =0.067, p=0.002) are significant independent predictors of the impairment score, along with their interaction (β =-0.005, p=0.004), while grammatical comprehension is not a significant independent predictor (β =0.11, p=0.19). The results of this analysis suggest that social cognition and executive functioning interact with one another and may play a role in the interpretation of indirect speech in bvFTD.

### 3.3 Neuroimaging Analyses

We also sought to determine the neuroanatomic basis of indirect speech act comprehension. More specifically, we examined regions of gray and white matter disease that may be related to impaired indirect speech act performance in patients with bvFTD. We note here that we focus solely on bvFTD patients in the following analyses because patients with MCI showed no impairment in the indirect condition, which is our experimental condition of interest.

We first contrasted cortical thickness in patients with bvFTD relative to an independent cohort of age-matched healthy controls. As illustrated in Figure 3 and summarized in Table 4, this reveals significantly reduced cortical thickness throughout the frontal and anterior temporal lobes bilaterally in bvFTD, with a peak in orbitofrontal cortex, consistent with disease diagnosis and previous structural imaging studies (Rascovsky et al., 2011; Möller et al., 2016).

**Table 4.** MNI Coordinates for Structural Imaging Results.

| Neuroanatomic Region (BA) | L/R | MNI Coordinates |  |  | t-stat | voxels |
| --- | --- | --- | --- | --- | --- | --- |
|  |  | x | y | z |  |  |
| A. Regions of cortical thinning in bvFTD relative to healthy controls |  |  |  |  |  |  |
| medial orbitofrontal cortex (11) | L | -2 | 46 | -20 | 6.75 | 39, 637 |
| middle temporal gyrus (12) | R | 60 | -12 | -24 | 6.43 | sub |
| orbitofrontal cortex (47) | R | 32 | 36 | -12 | 6.42 | sub |
| insula (13) | R | 40 | 2 | 2 | 6.42 | sub |
| medial orbitofrontal cortex (11) | R | 6 | 50 | -24 | 6.33 | sub |
| temporoparietal junction (39) | R | 40 | -54 | 24 | 6.29 | 79 |
| angular gyrus (39) | L | -42 | -60 | 32 | 4.19 | 33 |
| premotor cortex (6) | L | -46 | 2 | 34 | 4.77 | 27 |
| dorsolateral prefrontal cortex(9) | L | -22 | 36 | 26 | 3.25 | 12 |
| B. Regions of cortical thinning in bvFTD related to indirect impairment |  |  |  |  |  |  |
| inferior frontal gyrus (pars opercularis) (44) | R | 44 | 20 | 4 | 3.79 | 146 |
| precuneus (31) | R | 4 | -54 | 40 | 3.91 | 139 |
| posterior middle temporal gyrus (21) | L | -54 | -36 | 0 | 3.19 | 77 |
| premotor cortex (6) | L | -30 | -14 | 58 | 4.84 | 66 |
| premotor cortex (6) | R | 38 | -4 | 52 | 3.28 | 65 |
| medial prefrontal cortex (10) | L | -2 | 62 | -10 | 4.38 | 53 |
| inferior frontal gyrus (pars orbitalis) (47) | L | -38 | 24 | -2 | 3.5 | 50 |
| orbitofrontal cortex (11) | L | -22 | 34 | -20 | 3.48 | 49 |
| frontal pole (10) | L | -14 | 64 | -6 | 3.7 | 41 |
| dorsolateral prefrontal cortex (46) | L | -42 | 42 | 20 | 3.73 | 40 |
| posterior middle temporal gyrus (21) | L | -68 | -42 | -2 | 3.49 | 37 |
| dorsolateral prefrontal cortex (9) | L | -22 | 36 | 40 | 3.99 | 34 |
| precentral gyrus (4) | R | 28 | -24 | 64 | 2.75 | 32 |
| premotor cortex (6) | L | -26 | 4 | 54 | 4.51 | 31 |
| supplementary motor area (6) | L | -6 | -20 | 70 | 3.22 | 28 |
| dorsolateral prefrontal cortex (46) | L | -46 | 36 | 14 | 3.7 | 28 |
| supplementary motor area (6) | R | 4 | -20 | 72 | 3.05 | 21 |

Next, to relate patient deficits in indirect speech act comprehension to gray matter disease, we performed a regression analysis using the impairment score (indirect - direct) as a covariate. We find that greater relative impairment in the indirect condition is related to reduced cortical thickness in a largely left-lateralized cortical network, spanning frontal, temporal, and parietal regions. Significant clusters are observed within the classic peri-Sylvian language network, including left inferior frontal gyrus and posterior middle to superior temporal gyri, as well as right inferior frontal gyrus (pars opercularis). Additional effects are seen in regions that are more traditionally associated with social cognition, including medial prefrontal cortex, orbitofrontal cortex, and precuneus; or with executive function, including dorsolateral prefrontal cortex. Although unpredicted, we also see significant associations with premotor cortex, precentral gyrus, and supplementary motor areas, which have been previously implicated as part of the multiple-demand network and thought to play a role in broad domain-general functions (Fedorenko et al., 2013).

Our next set of analyses tested our hypothesis that three primary networks (language, social, and executive) are related to indirect speech comprehension by computing linear models using the mean cortical thickness score for each network as predictors for the impairment score. We did this by using a ROI-based approach across the whole-brain, rather than a voxel-wise approach. Using the network ROIs associated with language, social, and executive function defined in the Methods section, we find significant effects for each of our three networks, as shown in Figure 4. This effect is specific to these 3 networks and is not observed in the negative control, sensorimotor network.

**Figure 4.**
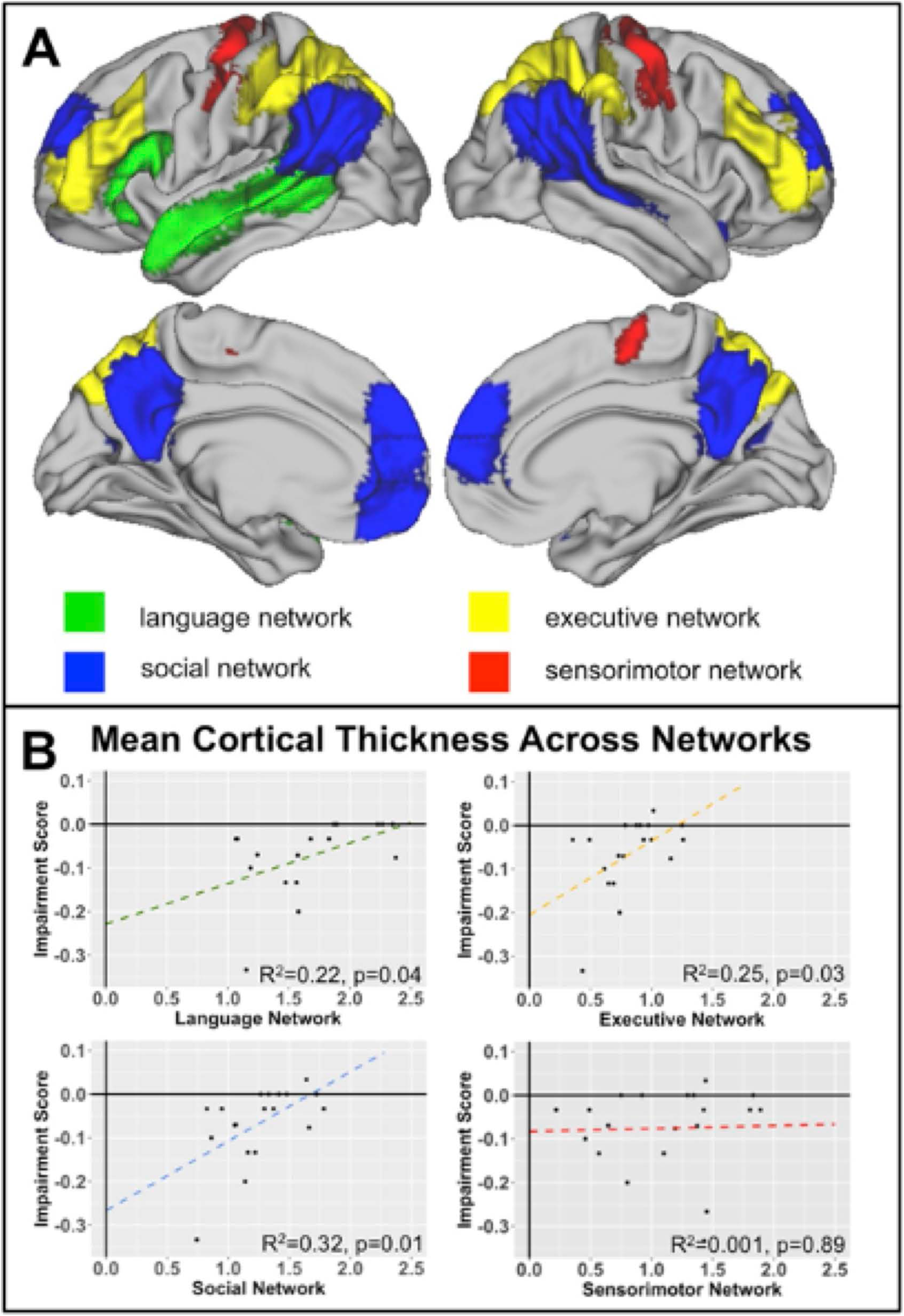
Network Analyses: Panel A: Network Key. Surface renderings of the brain showing each of the 4 network ROIs tested for their relationship with indirect speech processing: language network (green), social network (blue), executive network (yellow), and sensorimotor network (red). See text for a description of how each network was defined. Panel B: Network associations with Indirect comprehension impairment. Graphs plot the relationships between network cortical thickness in behavioral variant frontotemporal dementia (bvFTD) and indirect impairment score for language, executive, social, and sensorimotor networks. Note that the sensorimotor network is included as a negative control network to demonstrate specificity. See bottom right corner of each plot for R^2^ values.

Finally, while the majority of previous work on language comprehension has focused primarily on gray matter contributions to processing, we adopt a more connectionist approach here. Using a voxel-wise approach, we observe a significant change in FA in the following tracts within bvFTD: uncinate fasciculus, superior and inferior longitudinal fasciculi, and inferior fronto-occipital fasciculus. These are all long-range association tracts. We also observed disease in the corpus callosum, as well as white matter of the middle frontal and temporal gyri. We next examined which of these tracts are associated with the (indirect – direct) impairment score in bvFTD. As illustrated in Figure 5 and summarized in Table 5, we find significant effects for the superior longitudinal fasciculus (typically implicated in language processing), as well as the uncinate fasciculus (typically implicated in social-behavioral functioning), and inferior fronto-occipital fasciculus and frontal aslant.

**Table 5.**
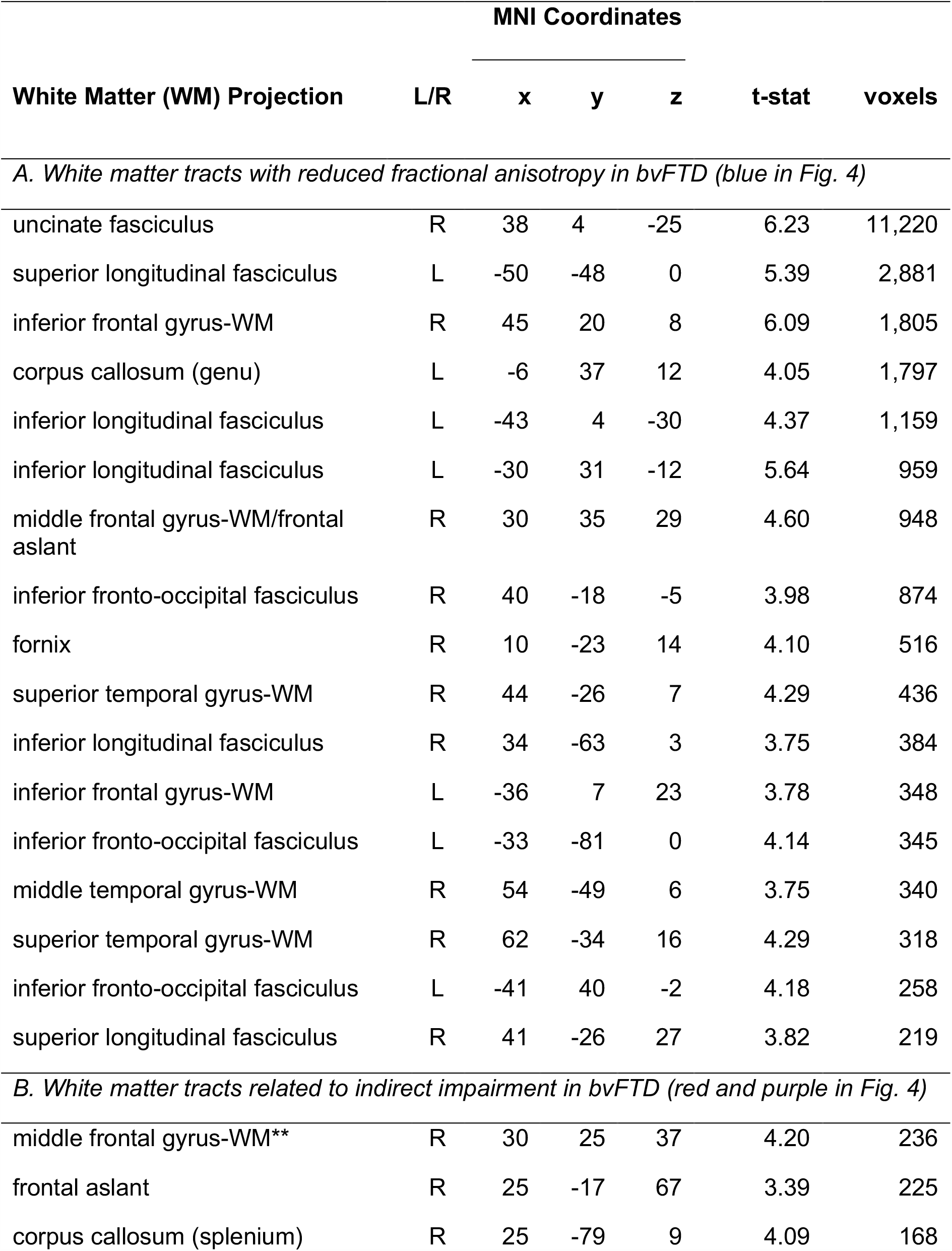
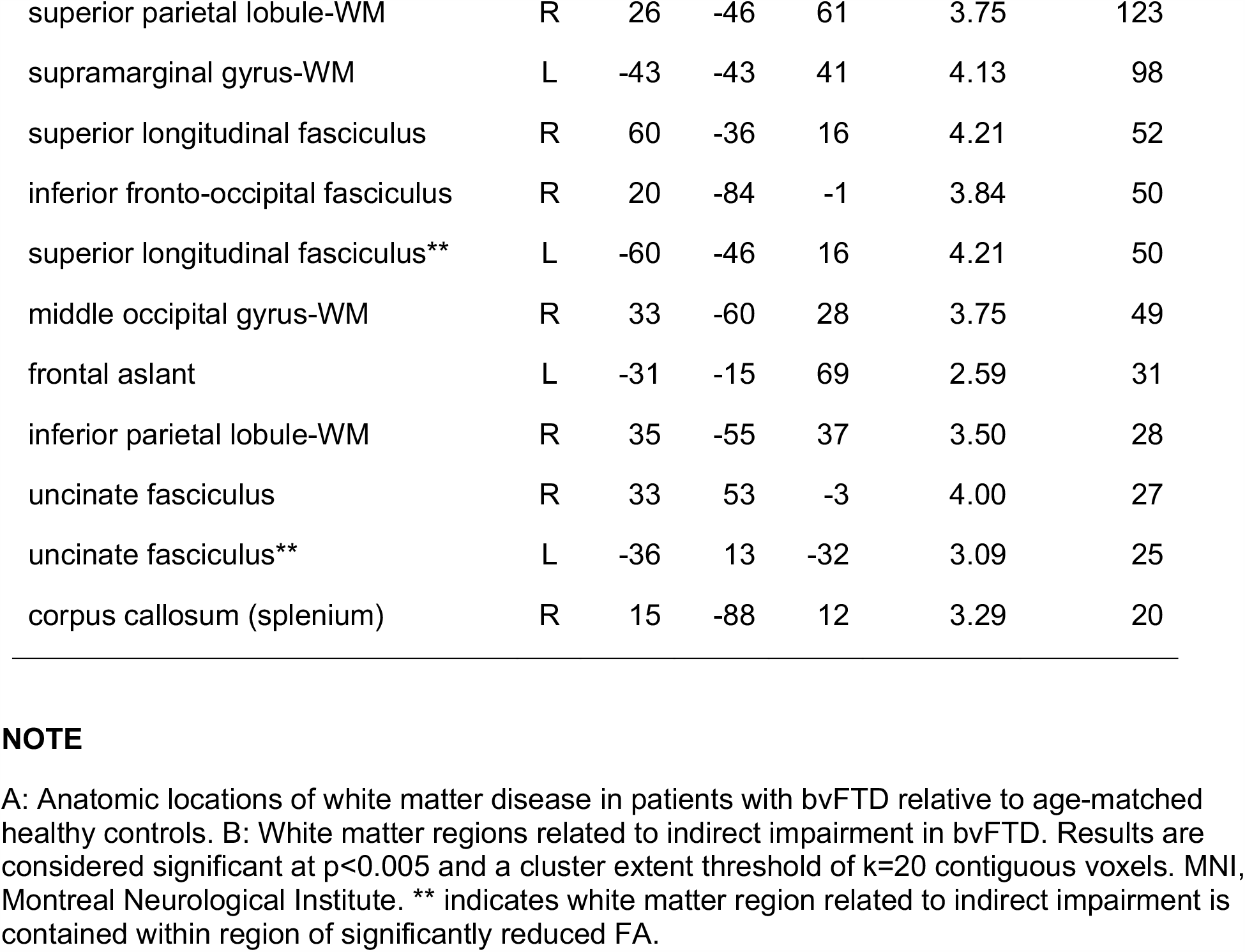
MNI Coordinates for Diffusion Tensor Imaging Results.

| White Matter (WM) Projection | L/R | MNI Coordinates |  |  | t-stat | voxels |
| --- | --- | --- | --- | --- | --- | --- |
|  |  | x | y | z |  |  |
| A. White matter tracts with reduced fractional anisotropy in bvFTD (blue in Fig. 4) |  |  |  |  |  |  |
| uncinate fasciculus | R | 38 | 4 | -25 | 6.23 | 11,220 |
| superior longitudinal fasciculus | L | -50 | -48 | 0 | 5.39 | 2,881 |
| inferior frontal gyrus-WM | R | 45 | 20 | 8 | 6.09 | 1,805 |
| corpus callosum (genu) | L | -6 | 37 | 12 | 4.05 | 1,797 |
| inferior longitudinal fasciculus | L | -43 | 4 | -30 | 4.37 | 1,159 |
| inferior longitudinal fasciculus | L | -30 | 31 | -12 | 5.64 | 959 |
| middle frontal gyrus-WM/frontal aslant | R | 30 | 35 | 29 | 4.60 | 948 |
| inferior fronto-occipital fasciculus | R | 40 | -18 | -5 | 3.98 | 874 |
| fornix | R | 10 | -23 | 14 | 4.10 | 516 |
| superior temporal gyrus-WM | R | 44 | -26 | 7 | 4.29 | 436 |
| inferior longitudinal fasciculus | R | 34 | -63 | 3 | 3.75 | 384 |
| inferior frontal gyrus-WM | L | -36 | 7 | 23 | 3.78 | 348 |
| inferior fronto-occipital fasciculus | L | -33 | -81 | 0 | 4.14 | 345 |
| middle temporal gyrus-WM | R | 54 | -49 | 6 | 3.75 | 340 |
| superior temporal gyrus-WM | R | 62 | -34 | 16 | 4.29 | 318 |
| inferior fronto-occipital fasciculus | L | -41 | 40 | -2 | 4.18 | 258 |
| superior longitudinal fasciculus | R | 41 | -26 | 27 | 3.82 | 219 |
| B. White matter tracts related to indirect impairment in bvFTD (red and purple in Fig. 4) |  |  |  |  |  |  |
| middle frontal gyrus-WM** | R | 30 | 25 | 37 | 4.20 | 236 |
| frontal aslant | R | 25 | -17 | 67 | 3.39 | 225 |
| corpus callosum (splenium) | R | 25 | -79 | 9 | 4.09 | 168 |
| superior parietal lobule-WM | R | 26 | -46 | 61 | 3.75 | 123 |
| supramarginal gyrus-WM | L | -43 | -43 | 41 | 4.13 | 98 |
| superior longitudinal fasciculus | R | 60 | -36 | 16 | 4.21 | 52 |
| inferior fronto-occipital fasciculus | R | 20 | -84 | -1 | 3.84 | 50 |
| superior longitudinal fasciculus** | L | -60 | -46 | 16 | 4.21 | 50 |
| middle occipital gyrus-WM | R | 33 | -60 | 28 | 3.75 | 49 |
| frontal aslant | L | -31 | -15 | 69 | 2.59 | 31 |
| inferior parietal lobule-WM | R | 35 | -55 | 37 | 3.50 | 28 |
| uncinate fasciculus | R | 33 | 53 | -3 | 4.00 | 27 |
| uncinate fasciculus** | L | -36 | 13 | -32 | 3.09 | 25 |
| corpus callosum (splenium) | R | 15 | -88 | 12 | 3.29 | 20 |

**Figure 5.**
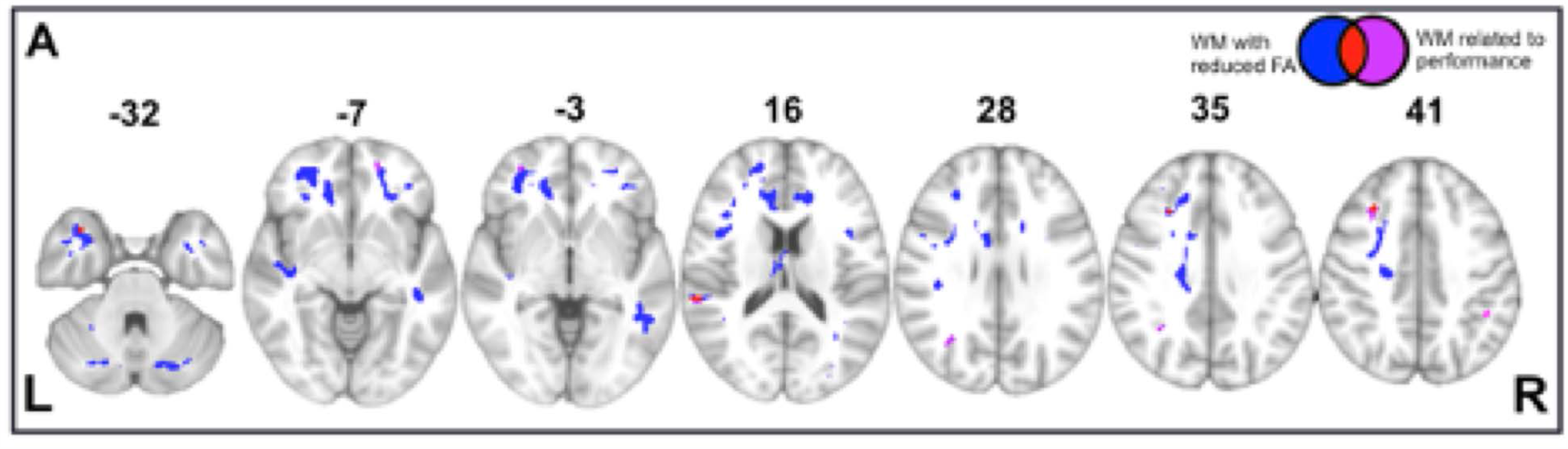
White Matter Imaging Results: Panel A: Axial slices showing regions of significantly reduced fractional anisotropy (FA) in white matter (WM) of behavioral variant frontotemporal dementia (bvFTD) patients relative to age-matched healthy controls (blue), regions of significantly reduced FA related to indirect impairment (red), and ancillary white matter regions (outside of blue regions of disease) also related to indirect impairment (violet). See key in upper right hand corner.

## 4 DISCUSSION

Most listeners are exceedingly adept at decoding a speaker’s intended meaning, despite the ambiguity inherent in much conversational speech. An unresolved question in neuroscience is how the brain accomplishes this feat. To address this issue, we study speech act processing in non-aphasic patients with bvFTD, and demonstrate that their comprehension is most impaired when a speaker’s intended meaning is communicated indirectly. This is related in part to neuropsychological measures of social and executive functioning and grammatical comprehension. Moreover, the observed impairment in indirect speech act comprehension is related to disease not only in the traditional language network (including left IFG and pMTG/STG), but also in two additional networks: the social brain network (including mPFC, OFC, and precuneus) and the executive network (including DLPFC, premotor cortex, and supplementary motor area), as well as the long-tract white matter projections that integrate these networks. Therefore, while traditional models of language highlight a left peri-Sylvian network, our observations are consistent with the hypothesis that the highly common but often overlooked inferential component of conversational speech is supported in part an extended language network that also incorporates frontal and parietal association cortices well beyond traditional left peri-Sylvian language regions. We discuss these findings and their implications below.

### 4.1 Inferential Demand Modulates Language Comprehension

Our primary objective was to examine how inferential demand—whether a speaker’s message is communicated directly or indirectly—modulates comprehension. Analyses of patient performance based on both accuracy and reaction time metrics suggests a deficit in indirect speech act comprehension in bvFTD. Findings of reduced accuracy extend the social-executive deficit in bvFTD to the language domain. We are unaware of other studies of indirect speech act comprehension in bvFTD, although clinical observations of schizophrenia, autism, and traumatic brain injury suggest difficulties with indirect speech exist in these populations (Champagne-Lavau and Stip, 2010; Johnson and Turkstra, 2012; Pastor-Cerezuela et al., 2018). Even for items where comprehension is preserved, moreover, we find significantly slowed performance. Consistent with our findings, previous studies have also reported that reaction time increases along with higher inferential demand (Ferstl and von Cramon, 2002; Kuperberg et al., 2006; Siebörger et al., 2007). Slowed processing can have considerable effects on real-world communication, as the gap between “turns at talk” is typically on the order of 200-250ms (Stivers et al., 2009; Levinson, 2016). In our data, bvFTD patients show a slowing effect of ∼600ms: such a processing lag would obviously impede the rapid switching that characterizes human conversation.

We further demonstrate that our effects are specific to bvFTD and not observable in brain-damaged controls with MCI. While evidence suggests that pragmatic deficits, including in proverb interpretation, exist in MCI (Leyhe et al., 2011; Cardoso et al., 2014), such findings may be a consequence of experimental confounds related to stimulus length or “frozen” meanings, and potentially associating findings with impaired episodic memory retrieval rather than impaired inferential processing. More work is needed to investigate this possibility.

Next, we examined the cognitive mechanisms that may contribute to indirect speech comprehension by evaluating the results of a neuropsychological battery. Results indicate that the observed deficit in bvFTD may be multifactorial in nature, as the indirect speech impairment (indirect – direct) score is related in part to language, social, and executive functioning, but not to episodic memory or visuospatial functioning.

Consider first executive function, which was assessed by Trailmaking. This finding aligns well with previous research showing a relationship between mental flexibility and pragmatic competence (Eslinger et al., 2007; Torralva et al., 2015). For example, Torralva et al. (2015) demonstrated that cognitive theory of mind and the ability to infer a speaker’s intention in a faux pas task is related to Trailmaking performance, although this finding is confounded in part by the lengthy nature of the stimulus items. While using a narrative helps establish context, as in these stimulus materials, this can also increase executive demands and introduce carry-over effects that make it difficult to dissociate inferential processing from task-related components of narrative processing, such as working memory demands needed to maintain narrative elements and track a character over time. Other results emphasize this potential confound by suggesting that working memory capacity may predict inference revision ability (Tompkins et al., 1994; Wright and Newhoff, 2002; Pérez et al., 2014). In our experiment, we suggest that bvFTD patients struggle to infer a speaker’s intention, perhaps related to difficulty switching from a literal to a pragmatic interpretation of utterance meaning. Our results are less likely to be confounded by working memory demands due to the brief stimulus items and the availability of written stimuli during the entire procedure. Indeed, although working memory is decreased in bvFTD (Kramer et al., 2003), we find little relationship between indirect speech act comprehension impairment and digit span. Future work using auditory stimuli should further investigate working memory contributions to language comprehension.

We also report a positive association between the indirect speech impairment score and performance on SNQ—a questionnaire assessing an individual’s ability to apply socially-dictated rules given different constraints (e.g. a conversation with a stranger versus a friend).One important social norm for conversational exchanges is Grice’s “Maxim of Relevance,” which states that an individual’s contribution to an ongoing exchange should always be pertinent and on-topic. bvFTD subjects may fail to appreciate this maxim due to degraded social knowledge, and thus they may judge indirect speech as irrelevant to the ongoing exchange and disregard it accordingly, ultimately resulting in impaired comprehension, as observed here. Task demands also depend in part on meta-judgments, that is, the patient’s judgment of whether a reply means “yes” or “no.” This kind of judgment in a controlled experimental context may differ from judgments in a real-world context where there is also additional contextual support for responding to an indirect request. While indirect responses to some requests may be overlearned (e.g. Question: “Do you have the time?” Response: “Noontime.”) relative to a direct response (Question: “Do you have the time?” Response: “Yes.”), the clinical impression is that questions eliciting an indirect response tend to yield a direct or literal response from bvFTD patients more often than would ordinarily be expected. Additional work is needed in a real-world setting to gauge the extent of an indirect speech act comprehension deficit in bvFTD.

Multiple regression analysis confirms the role that executive function and social cognition play in impairment. The final model (Impairment Score ∼ Grammatical Comprehension + Trailmaking * SNQ) also suggests that social and executive deficits are not independent, but rather interact. This result has implications for an ongoing debate in the bvFTD literature concerning the relationship between social cognition and executive function (Lough et al., 2001; Eslinger et al., 2007; Le Bouc et al., 2012; Bertoux et al., 2016).

Although patients with bvFTD are grossly non-aphasic according to clinician assessments of speech, their indirect impairment is associated in part with a language measure—grammatical comprehension. In this case, grammatical comprehension was assessed by comparing sentence-picture matching for object-relative compared to subject-relative sentences. We note here that the comprehension of object-relative phrases is known to be more difficult than subject-relative phrases in both healthy adults and patients with bvFTD (Charles et al., 2014; Demberg and Sayeed, 2016). This positive association may be related in part to the mental manipulation of linguistic materials that may contribute to comprehension of both object-related phrases and indirect speech acts. We also note here that the relationship between grammatical comprehension and indirect speech act comprehension impairment is lessened when concomitant deficits in social cognition and executive function are taken into account in our three-factor regression model. Additional work is needed to examine the nature of deficits in grammatical comprehension in bvFTD, and the potential contribution of such a deficit to the comprehension of indirect speech acts in these patients.

While we used carefully constructed and well-matched experimental materials to assess comprehension of indirect relative to direct speech acts, one shortcoming of our study is that performance depends on patients’ judgments of speech acts rather than their difficulty engaging in actual speech acts during day-to-day natural discourse. To mitigate this shortcoming, we related patients’ indirect speech act performance to caregivers’ judgments of communicative efficacy. We find that difficulty with indirect speech act comprehension is related to caregivers’ perception of impoverished day-to-day conversational ability. Ultimately, our goal is to help improve daily discourse in bvFTD in order to advance a patient’s quality of life and minimize health-related risks associated with limitations in conversational exchanges that frequently involve indirect speech acts. Consider a situation where a caregiver asks a loved one “Do you need me?”, and the patient with bvFTD responds “Yes.” A direct reply to this request, when an indirect response to a question is wished, for example, does not indicate that the patient is feeling chest pain as might be experienced in a myocardial infarct. Instead, the caregiver may be obliged in this situation to make an inference based on the patient’s facial expression of pain. It is exceedingly common that a caregiver infers a bvFTD patient’s needs based on information other than that most often available from a verbal response. Additional work is needed to confirm the consequences of impaired indirect speech act comprehension in natural, day-to-day conversation in bvFTD.

### 4.2 An Extra-Sylvian Network for Speech Act Comprehension

Although neurobiological models of language centered on left peri-Sylvian regions have been foundational in studies of human brain functioning, these models remain limited in their external validity and generalizability to real-world contexts (Hasson et al., 2018). Here, we examine cortical thinning and fractional anisotropy in patients with bvFTD, and begin to build a large-scale, multimodal language network that can potentially account for some common aspects of real-world discourse such as indirect speech act comprehension.

To date, only a limited number of studies has examined the neural basis of indirect reply comprehension (Shibata et al., 2011; Basnáková et al., 2013; Jang et al., 2013; Feng et al., 2017). While these fMRI studies offer preliminary evidence for the role of non-language regions including mPFC, TPJ, and precuneus in discourse processing, there are some caveats to keep in mind. For example, several studies used experimental tasks that involve reading a brief narrative followed by an exchange between speakers. Using narratives to establish context can increase executive demands and introduce carry-over effects that make it difficult to dissociate inferential processing from other task-related components of narrative processing—including tracking a character over time, processing event structure, maintaining narrative elements in working memory, and more. To help manage this potential confound, we designed a novel question-answer paradigm that manipulated inferential demand while simultaneously minimizing task-related resource demands and controlling for linguistic variation across stimuli. fMRI studies also are correlative in nature and may not fully control for all task-related demands, prompting us to focus on a “lesion-based” model to examine cerebral-cognitive functioning.

As our paradigm was language-based, we did observe significant effects in the IFG and the posterior MTG/STG. These areas, initially proposed by the WLG model and later confirmed, constitute the primary nodes of the classic language network (Binder et al., 1997; Price CJ, 2000). In our study, we point out that these peri-Sylvian regions are related to patient impairment in the indirect condition *over and above* that found in the direct condition. Therefore, our results support a role for left peri-Sylvian regions not only in lexical, semantic, and syntactic processing, but also in higher-level selection and global integration that may be required in discourse, as suggested previously (Hagoort, 2005). Previous fMRI studies of indirect speech and causal inferencing make similar arguments (Mason and Just, 2004; Eviatar and Just, 2006). We also observe an effect in the right IFG, which is consistent with the dynamic spillover hypothesis described by Prat and colleagues (Prat et al., 2011). According to this model, activity in the right hemisphere is more likely to be invoked when 1) readers are less skilled, and 2) passage difficulty is harder.

We now know that language processing also extends beyond peri-Sylvian regions (Ferstl et al., 2008; Fedorenko and Thompson-Schill, 2014; Hagoort, 2014). We report here that extra-Sylvian regions, including the orbitofrontal, medial prefrontal, dorsolateral prefrontal cortices, as well as precuneus and premotor and supplementary motor regions, are related to indirect speech processing in bvFTD. These are regions encompassed by social and executive networks of the brain. Importantly, these findings are relatively selective, as we find no evidence of other network involvement (e.g. sensorimotor network).

Consider first mPFC and precuneus, which are both related to a social brain network commonly associated with “theory of mind” (Saxe and Kanwisher, 2003; Frith and Frith, 2012; Dufour et al., 2013; Healey and Grossman, 2018). While mPFC is traditionally associated with perspective-taking and the ability to make inferences about conspecifics, recent research also suggests that a ventral portion of mPFC, similar to the cluster observed here, plays a role in scene construction and situational processing (Lieberman et al., 2019). In the case of indirect speech acts, mPFC may help generate a “schema” or “situation model” that guides interpretation of ambiguous stimuli and events. The precuneus may play a similar role. One of the brain’s most globally connected areas, the precuneus is traditionally associated with a diverse set of cognitive functions including visuospatial processing, episodic memory (Shallice et al., 1994), and mental imagery (Hassabis et al., 2007; Johnson et al., 2007). Newer work, however, has demonstrated that the precuneus also plays a role in self-referential processing and first-person perspective-taking, as well as situation model-building and the retrieval of contextual associations from internal stores (Lundstrom et al., 2005; Cavanna and Trimble, 2006; Binder et al., 2009; Mashal et al., 2014; Herold et al., 2016). Taken together, the relationship of mPFC and precuneus to indirect speech impairment suggests that indirect reply comprehension requires listeners to 1) adopt the speaker’s perspective, and 2) integrate contextual information into some kind of mental model. Finally, we also observed an effect in orbitofrontal cortex, which is sometimes included in the social brain network. Like mPFC and precuneus, some studies implicate OFC in theory of mind, in addition to tasks involving reversal learning, set-shifting, and affective decision-making (Rolls, 2004; Sabbagh, 2004; Badre and Wagner, 2006).

DLPFC, on the other hand, is often considered to be part of a “multiple-demand” network commonly linked to domain-general, executive control processes that may be involved in language and other behaviors (Novais-Santos et al., 2007; Duncan, 2010; Fedorenko et al., 2013). These regions are often defined by contrasting two task conditions that vary in difficulty (e.g. verbal working memory tasks with 4 digits versus 8 digits), mirroring our indirect-direct contrast. It is important to note that these “harder” tasks may not only require additional computational resources, but could also invoke strategic reasoning processes mediated by DLPFC (Yoshida et al., 2010; Yamagishi et al., 2016). Finally, with well-documented roles in working memory and selection (Petrides, 2005; Badre, 2008), DLPFC is also implicated in the Memory-Unification-Control model of language (Hagoort, 2013), serving as the “control” component and mediating processes such as turn-taking and the selection of contextually-appropriate meanings. The motor-associated regions we observed, including premotor cortex, precentral gyrus, and supplementary motor area, have also been hypothesized to belong to this same network as DLPFC (Fedorenko et al., 2013). While additional work is needed to determine more precisely the cortical constituents underlying the social and executive components of language, our extended neurobiological model of language proposes that a neural network supporting common elements of discourse such as indirect speech act comprehension incorporating these brain regions associated with executive and social functioning.

Other materials have been used to study inferential demands in language, but may be associated with confounds that can limit interpretation. For example, recent work using fMRI (Paunov et al., 2019) has demonstrated that story comprehension elicits synchronized network activity not only in traditional language-associated regions, but also in social regions including mPFC, TPJ, and precuneus. Similar results have been reported elsewhere (Xu et al., 2005; Mar, 2011; AbdulSabur et al., 2014). A third, fronto-parietal network associated with executive control, possibly related to working memory, has also been implicated in story comprehension (Raposo and Marques, 2013; Smirnov et al., 2014; Mineroff et al., 2018; Aboud et al., 2019; Paunov et al., 2019). Although these results are promising, narratives are inherently long, which makes them difficult to control experimentally and overly dependent on task-related executive resources.

Another common approach to examining the inferential component of discourse has been the study of non-literal or figurative language, including sarcasm, irony, metaphors, idioms, and proverbs (see Rapp et al., 2012 for a comprehensive review). This body of work also implicates social and executive components in the comprehension of pragmatic language (Wakusawa et al., 2007; Bohrn et al., 2012; Uchiyama et al., 2012; Iskandar and Baird, 2014; Obert et al., 2016; Filik et al., 2019). While informative, these materials are often subject to confounds related to varying degrees of familiarity and emotional valence, among others (Nippold and Haq, 1996; Schmidt and Seger, 2009; Ziv et al., 2011; Kaiser et al., 2013). To minimize confounds associated with lengthy, contextualizing narratives, differential familiarity, and emotional valence, we opted to study the neural basis for inferential reasoning in discourse by studying indirect speech acts, where we found that comprehension encompasses a core, left peri-Sylvian language network as well as cortical regions often encompassed by executive controls and social networks.

### 4.3 White Matter Correlates of Speech Act Comprehension

Recent work has paid increasing attention to white matter connectivity. While the traditional WLG model of language focuses primarily on the arcuate fasciculus-- a component of the superior longitudinal fasciculus connecting IFG and STG, newer work has identified pathways that not only interconnect peri-Sylvian regions, but also connect these regions to extra-Sylvian regions (Catani et al., 2005; Dick and Tremblay, 2012). Analogous to the visual system, these pathways may be divided into dorsal and ventral streams. One characterization implicates the dorsal stream as broadly involved in auditory-motor integration and the ventral stream in mapping form to meaning (Hickok and Poeppel, 2004; Saur et al., 2008). Using voxel-based fractional anisotropy analyses, we find evidence implicating tracts in both dorsal and ventral streams in indirect speech act comprehension. This includes the uncinate and inferior fronto-occipital fasciculi in the ventral stream, and the superior longitudinal fasciculus and frontal aslant in the dorsal stream. The frontal aslant, in particular, is a recently discovered tract implicated in both speech and language (on the left) and executive function (on the right) (Varriano et al., 2018; Dick et al., 2019). The frontal aslant, which is thought to project between the IFG and the supplementary motor areas, has previously been implicated in verbal fluency deficits in aphasic variants of frontotemporal dementia known as primary progressive aphasia (Catani et al., 2013). The uncinate fasciculus, carrying projections between orbitofrontal cortex and anterior temporal regions, has also gained more attention recently as a white matter tract mediating the interaction of social and language networks. For example, damage to the uncinate fasciculus is bvFTD is associated not only with a bvFTD diagnosis, but also with deficits in non-literal language comprehension including sarcasm and irony (Agosta et al., 2012; Downey et al., 2015). Thus, we propose that cortical components of our extended language network are integrated by white matter projections in both dorsal and ventral projection streams.

### 4.4 Conclusions

Strengths of our study include the novel task design with carefully matched direct and indirect conditions and minimal task-related demands, observation of a significant indirect language impairment in a non-aphasic brain-damaged cohort with selective social and executive deficits, and robust association of these deficits with an anatomic network implicating language, social, and executive networks. Nevertheless, several caveats should be kept in mind when interpreting our results. Although we tested a relatively large bvFTD cohort and demonstrated specificity with a brain-damaged control group, our cohort is small, patients are not pathologically confirmed, and generalizability is limited to the mild-moderate disease stage. Second, while we confirmed an indirect speech impairment with reaction time data, we report ceiling effects for accuracy in our control subjects, thereby limiting examination of individual differences associated with aging. To differentiate the functions of nodes within the extended language network, future experimental studies should contrast different types of indirect speech, including indirect requests (which have a motor component) and “face-saving” replies (which have an affective component), and assess indirect speech acts in natural discourse. While correlative, it would be valuable to confirm our anatomic observations in healthy young adults using fMRI, and to confirm a causal association for these anatomic observations in bvFTD using longitudinal studies.

The findings discussed here also have meaningful clinical implications. Communication difficulties can compromise social interactions, and in turn, diminish safety, interpersonal relationships and overall well-being. We find that impaired indirect speech is related to communicative efficacy, which is a crucial element of patient safety and quality of life. Accordingly, language deficits may be a target for intervention in bvFTD. Our data also have implications for “best-practice” communication strategies used by patient caregivers: to optimize successful communication, language should be as direct as possible.

With these caveats in mind, we conclude that patients with bvFTD struggle to make the pragmatic inferences necessary to support comprehension of indirect replies, a very common but understudied example of conversational discourse that depends on inferencing. This is due in part to social-executive deficits and degradation of a multimodal, extra-Sylvian network supporting natural, daily language use. More specifically, our findings emphasize the extension of the brain’s traditional language network beyond left peri-Sylvian regions into additional frontal and parietal regions that may be incorporated into a real-world language network that supports everyday discourse. We conclude by emphasizing the importance of studying language in context—the way in which we use it in everyday life. Indeed, it is only when we study language in this way—as a means of communication—that we can begin to characterize the full extent of its neurobiologic foundations.

## Acknowledgements

We thank the radiographers at the Hospital of the University of Pennsylvania for their assistance with data collection and our patients and volunteers for their continued participation.

## Notes

### Competing Interest Statement

The authors have declared no competing interest.

### Funding Statement

This work was supported in part by grants from NIH (AG066597, AG017586,AG052943, AG054519, NS101863), the Wyncote Foundation, and an anonymous
donor.

### Author Declarations

All subjects were recruited from the Penn Frontotemporal Degeneration Center and gave informed consent according to a protocol approved by the Institutional Review Board at the University of Pennsylvania.

